# Label-based meta-analysis of functional brain dysconnectivity across mood and psychotic disorders

**DOI:** 10.1101/2022.09.27.22280420

**Authors:** Stéphanie Grot, Salima Smine, Stéphane Potvin, Maëliss Darcey, Vilena Pavlov, Sarah Genon, Hien Nguyen, Pierre Orban

**Author notes:** co-first authors (Stéphanie Grot, Salima Smine). correspondence Address correspondence to Pierre Orban, Ph.D., Centre de recherche de l’Institut universitaire en santé mentale de Montréal, 7331 rue Hochelaga, Montréal, QC, H1N 3V2, Canada.

## Abstract

**BACKGROUND:** Psychiatric diseases are increasingly conceptualized as brain network disorders. Hundreds of resting-state functional magnetic resonance imaging (rsfMRI) studies have revealed patterns of functional brain dysconnectivity in disorders such as major depression disorder (MDD), bipolar disorder (BD) and schizophrenia (SZ). Although these disorders have been mostly studied in isolation, there is mounting evidence of shared neurobiological alterations across disorders.

**METHODS:** To uncover the nature of the relatedness between these psychiatric disorders, we conducted an innovative meta-analysis of past functional brain dysconnectivity findings obtained separately in MDD, BD and SZ. Rather than relying on a classical coordinate-based approach at the voxel level, our procedure extracted relevant neuroanatomical labels from text data and reported findings at the whole brain network level. Data were drawn from 428 rsfMRI studies investigating MDD (158 studies, 7429 patients / 7414 controls), BD (81 studies, 3330 patients / 4096 patients) and/or SZ (223 studies, 11168 patients / 11754 controls). Permutation testing revealed commonalities and specificities in hypoconnectivity and hyperconnectivity patterns across disorders.

**RESULTS:** Among 78 connections within or between 12 cortico-subcortical networks, hypoconnectivity and hyperconnectivity patterns of higher-order cognitive (default-mode, fronto-parietal, cingulo-opercular) networks were similarly observed across the 3 disorders. By contrast, dysconnectivity of lower-order (somatomotor, visual, auditory) networks in some cases differed between disorders, notably dissociating SZ from BD and MDD.

**CONCLUSIONS:** Our label-based meta-analytic approach allowed a comprehensive inclusion of prior studies. Findings suggest that functional brain dysconnectivity of higher-order cognitive networks is largely transdiagnostic in nature while that of lower-order networks may best discriminate mood and psychotic disorders, thus emphasizing the relevance of motor and sensory networks to psychiatric neuroscience.

## INTRODUCTION

The advent of functional magnetic resonance imaging (fMRI) 3 decades ago has greatly facilitated the investigation of neurobiological alterations in major psychiatric disorders such as major depressive disorder (MDD), bipolar disorder (BD) and schizophrenia (SZ) (1–3). The primary focus of psychiatric fMRI was the localisation of segregated brain regions with abnormal activation during performance of various cognitive tasks (4). A major development has then been the characterization of aberrant functional brain integration in mental disorders, which are now increasingly conceptualized as brain network disorders (5–8). This endeavor has been greatly promoted by relying on resting-state fMRI, with spontaneous slow fluctuations in brain activity defining large-scale networks of brain areas with correlated activity (9). Such intrinsic functional brain networks recapitulate with good correspondence the repertoire of brain regions co-activated during various behaviors (10). Current models of functional brain dysconnectivity in mental illness most consistently highlight abnormalities within and between 3 higher-order functional brain networks - noting their naming varies across the literature (11): the default-mode (medial prefrontal cortex, posterior cingulate cortex, posterior inferior parietal lobule), fronto-parietal (middle frontal gyrus, anterior inferior parietal lobule) and cingulo-opercular (anterior insula and anterior midcingulate cortex) networks (5,7,8,12–14).

As is the case for task-based activation studies, most resting-state connectivity research in mental illness has relied on case-control designs looking at psychiatric disorders in isolation. Consequently, findings from individual studies may attribute undue specificity of brain dysfunction patterns to one disorder or another. Indeed, there is growing acknowledgment that clinical (15), neurobiological (16) and genetic (17,18) boundaries between psychiatric disorders are blurrier than postulated by traditional classifications of mental illness such as the DSM-5. For instance, it is now clear that there is a greater continuum than once thought between mood and psychotic disorders, encompassing MDD, BD and SZ (19–23). Transdiagnostic research is thus needed to uncover the nature of the relatedness between psychiatric disorders, explaining their high comorbidity and facilitating the discovery of improved treatments (5,16,24,25). From a dimensional perspective, a common psychopathological factor shared among disorders has been associated with particular functional brain dysconnectivity patterns that transcend psychiatric diagnoses (26–30). Alternatively, direct comparative analysis of multiple diagnostic categories has allowed to evidence both commonalities and specificities in functional brain connectivity alterations (31–37). However, while findings that emerge from this burgeoning field of research are promising, original transdiagnostic studies are still scarce to date. Hence, there is value in conducting transdiagnostic meta-analyses contrasting various psychiatric disorders that were not investigated in the same original studies.

To date, transdiagnostic meta-analyses of both task-based (4,38–41) and resting-state (13,42,43) fMRI findings in neuropsychiatric disorders have mostly relied on coordinate-based approaches (44–46). When applied to case-control studies, methods such as activation likelihood estimation (ALE, (47)), multilevel kernel density analysis (MKDA, (48)) or signed differential mapping (SDM, (49)) quantitatively assess the spatial convergence of results from primary studies based on stereotactic coordinates of peak statistical differences between patients and controls. While coordinate-based meta-analysis has proven successful in unravelling consistent patterns across prior findings from activation studies, it is not best suited for synthesizing connectivity results. First, stereotactic coordinates are almost universally reported in activation studies (50) but much less so in the connectivity literature, among other reasons because analyses are often conducted at spatial resolutions other than the voxel level. Second, the prerequisite of a similar search coverage across studies (51) is met in most activation (most commonly the whole brain) but only few connectivity studies, in which region-of-interest analyses have long been the rule rather than the exception. Consequently, meta-analyses have often been restricted to including only studies with seeds falling within a few seed networks of interest (12–14,52,53), typically focussing on large, higher-order networks such as the default-mode, fronto-parietal, cingulo-opercular networks, and thereby ignoring less commonly explored networks (e.g., visual and somatomotor lower-order networks). To address these limitations, we conducted an innovative label-based meta-analysis (45,46,54,55) aimed at revealing commonalities and specificities in hypoconnectivity and hyperconnectivity patterns across MDD, BD and SZ. Our meta-analytical approach first avoids excluding studies that did not report coordinates by extracting findings from prior studies based on text data. In addition, it unravels consistent dysconnectivity patterns across both lower- and higher-order distributed networks covering the whole brain while controlling for unequal search coverage across studies.

## METHODS AND MATERIALS

### Study selection

A literature search was conducted in the PubMed database up to October12, 2021 in accordance with the Preferred Reporting Items for Systematic Reviews and Meta-Analyses (PRISMA) guidelines (http://www.prisma-statement.org) (see flowchart in Supplemental Figure S1). The search terms were: (“depression” OR “depressive” OR “bipolar” OR “schizophrenia” OR “psychosis”) AND (“functional magnetic resonance imaging” OR “fMRI”) AND (“rest” OR “resting”) AND “connectivity”. Original studies using resting-state functional magnetic resonance imaging to characterize functional brain dysconnectivity in psychiatric patients with an explicit diagnosis of major depressive disorder (MDD), bipolar disorder (BD) or schizophrenia (SZ) were eligible for inclusion. Noteworthy, studies were included regardless of whether stereotactic coordinates reflecting the peak locations of significant group differences were reported. Exclusion criteria were as follows: (1) not in English; (2) no direct comparison of patients with a healthy control group; (3) functional brain connectivity investigated through approaches other than the seed-based voxel-wise (SBVW), seed-to-region (STR), network-based connectome-wide (NBCW), independent component analysis (ICA) with dual regression, voxel-mirrored homotopic connectivity (VMHC), regional homogeneity (ReHo), and amplitude of low-frequency fluctuation (ALFF) methods (Supplemental Table S1); (4) no adequate correction for multiple comparisons; (5) entirely overlapping sample with identical search coverage reported in another publication.

### Data extraction

#### Label-based meta-analysis

Our meta-analytical method is based on the systematic and principled extraction of neuroanatomical terms describing which functional brain connections were investigated or were evidenced as significantly impaired in psychiatric patients compared to healthy controls. Text from all paper sections (abstract, introduction, methods, results, discussion, as well as figure legends and tables) was mined by experts in macroneuroanatomy. Our approach dealt with the following confounds: first, there are large variations in search coverage from one paper to another - a minority of papers considers pairwise connectivity for all brain regions, most of them instead focus on a small part of whole-brain connectivity; second, the spatial granularity at which connectivity is explored varies drastically across papers – from voxels to regions to networks; third, there is significant variability in the neuroanatomical nomenclature used in the literature.

We implemented a two-step procedure that first manually transcribed the gathered neuroanatomical information at the original level of description (region, network, whole brain), then translated it with reference to a single common network-level framework. We separately coded connections that were the object of a statistical test (*tested* connections) and those that showed a statistically significant (p < 0.05 after correction for multiple comparison) alteration (*impaired* connections). In the latter case, we further differentiated *hypoconnected* from *hyperconnected* connections. We however did not distinguish between enhanced and weakened connectivity patterns, as this distinction is seldomly made in the literature. Thus, hypoconnectivity may indicate either larger negative or reduced positive connectivity while hyperconnectivity may refer to either larger positive or reduced negative connectivity.

#### Two-step extraction

For each paper, we extracted pairs of neuroanatomical terms describing functional brain connections. Because distinct terms may refer to similar brain areas or networks, we first manually transcribed the terms used in original papers with best fits from a limited set of labelling schemes covering the entire spatial granularity range. Brain regions were labelled based on the Automated Anatomical Atlas (AAL, 116 regions, (56), AAL3 (170 regions, (57)) or Brodmann atlas (48 regions, WFU PickAtlas software, (58); distributed brain networks were labelled based on the Cole-Anticevic Brain Network Partition (CAB-NP, 12 networks, (59)); and larger brain components such as lobes or the entire hemisphere were defined using the TD atlas (WFU PickAtlas software, (58)). The whole gray matter was defined by a mask including all regions from the AAL3 atlas (57). For each type of contrast (MDD vs. HC, BD vs. HC, SZ vs. HC) found in a study and each type of connection (tested, hypoconnected, hyperconnected), comprehensive transcription of the relevant neuroanatomical information was accomplished with as few pairs of labels as possible.

The second step involved translating, in an automated manner, the labels obtained at various spatial resolutions into a single common large-scale network space, the 12-network CAB-NP (59) (see Supplemental Methods for a secondary analysis at the region level). This functional brain atlas covers the whole brain, with many networks spanning both the cortex, basal ganglia and cerebellum (Figure 1). Higher-order cognitive networks (default-mode, frontoparietal, dorsal attentional, cingulo-opercular) are dissociated from lower-order networks (primary and secondary visual, somatomotor, auditory) as well as from language and ventral (orbito-affective, ventral and posterior multimodal) networks. Because the original cortical parcels of the CAB-NP are surface-based, we created a publicly available volumetric version of the atlas for the whole brain. Of note, all networks are not of equal size (Supplemental Table S2), with 3 higher-order networks (default-mode, frontoparietal and cingulo-opercular networks) together amounting for over 50% of the total atlas volume. Brain regions corresponding to manually extracted labels were automatically assigned to the large-scale network with which they maximally overlapped, and region labels were translated into the network space accordingly. For each study, separately for each type of contrast available (MDD vs. HC, BD vs. HC, SZ vs. HC) and each type of connection (tested, hypoconnected, hyperconnected), we then coded the absence (0) or presence (1) of pairs of network labels defining each of the 12 within-network and 66 between-network connections.

**Figure 1.**
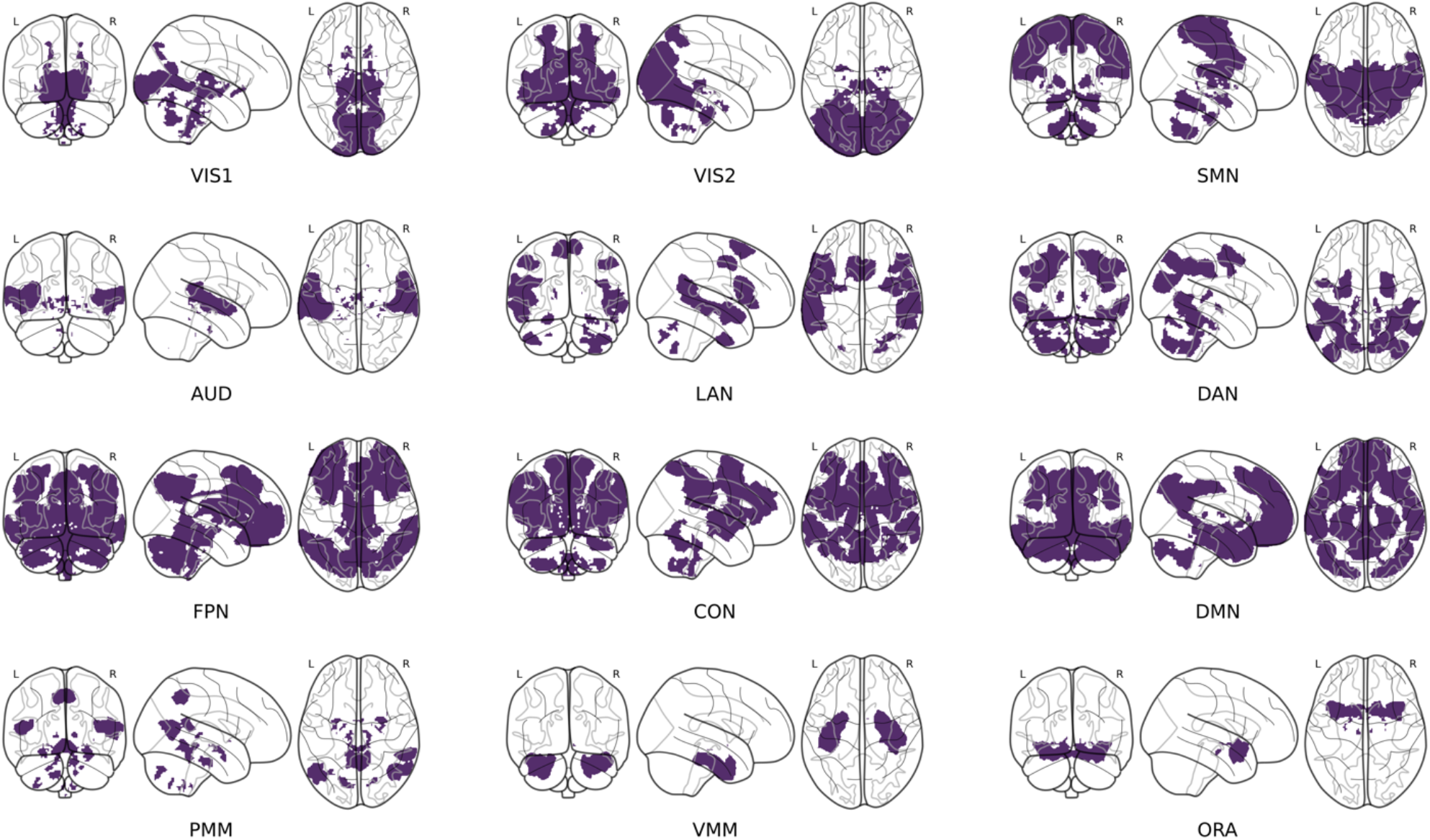
The Cole-Anticevic Brain Network Partition (CAB-NP) (59) was used as a reference space to report meta-analytic findings at the large-scale brain network level. This functional brain parcellation includes 12 cortico-subcortical distributed networks, here displayed on coronal, sagittal and axial views of glass brain representations. VIS1, primary visual network; VIS2, secondary visual network; SMN, sensorimotor network; AUD, auditory network; LAN, language network; DAN, dorsal attentional network; FPN, fronto-parietal network; CON, cingulo-opercular network; DMN, default-mode network; PMN, posterior multimodal network; VMN, ventral multimodal network; ORA, orbito-affective network.

#### Data extraction reliability

In addition to the main labels extraction for all 428 studies (SG1), separate raters (SP, MD, VP) together independently re-extracted labels for a subset of 100 studies. These latter studies were pseudo-randomly selected to ensure good representativity of the different types of studies (diagnostic group, connectivity method), in proportions similar to those found in the entire set (Figure 2). The similarity of the main and confirmatory network labels extractions for this subset of studies was computed with Dice similarity coefficients for binary matrices coding for the presence or absence of connection being tested or evidenced as impaired, over all studies and across all brain network connections defined by the CAB-NP. The similarity of labels extractions was quantified separately for the connections being tested and the connections being reported as impaired.

**Figure 2.**
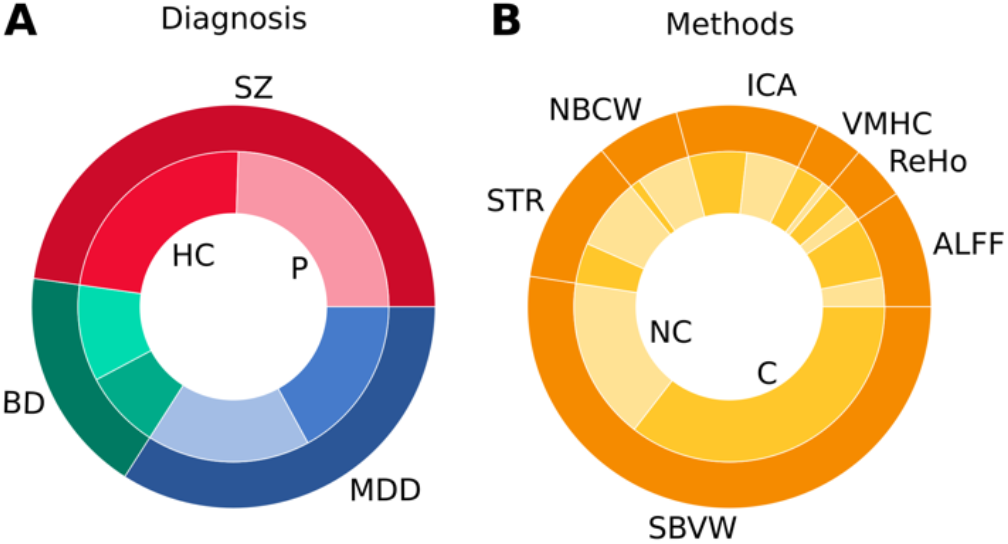
Proportions of the 428 studies that reported dysconnectivity effects for each psychiatric disorder (left, outer circle) and the respective proportions of patients (lighter color) and controls (darker color) for each diagnosis (left, inner circle). MDD, major depressive disorder; BP, bipolar disorder; SZ, schizophrenia; HC, healthy controls; P, patients. Proportions of the type of methodology used to characterize functional brain connectivity (right, outer circle), and the respective proportions of studies that reported stereotactic coordinates of peak effects (darker color) and did not (lighter color) for each type of methods (right, inner circle). SBVW, seed-based voxel-wise; STR, seed-to-region; NBCW, network-based connectome-wide; ICA, independent component analysis; VMHC, voxel mirrored homotopic connectivity; ReHo, regional homogeneity; ALFF, amplitude of low frequency fluctuations; C, with coordinates; NC, without coordinates. See Supplemental Methods for details.

### Statistical analysis

Permutation tests were conducted on one or more of 9 two-way binary tables indicating for each of the 78 connections whether it was tested, hypoconnected, hyperconnected (1) or not (0) in each of the studies looking at MDD, BD or SZ, respectively. Permutation tests (k = 100,000) investigated effects that were directly tested in original studies (hypoconnectivity or hyperconnectivity relative to HC in either of the 3 disorders) or not directly tested (hypoconnectivity vs. hyperconnectivity in either of the 3 disorders as well as pairwise comparisons between disorders for either hypoconnectivity or hyperconnectivity). Mathematical formulations are provided in the Supplemental Methods.

To test for hypoconnectivity (or hyperconnectivity) of each pair of networks (connection), we used a permutation testing approach whereupon a sample proportion test statistic (computed as a ratio of observed reported effects over baseline of whether the pair of networks in question was tested or not) was used. These ratios had numerators equal to the sum of number of studies that observed a significant level of either hypoconnectivity (or hyperconnectivity) for each pair of networks of interest, and had denominators equal to the number of studies that tested the pair of networks of interest as a baseline. We then sampled from a subset of the permutations applied to the observed outcomes of each study to randomize whether each observed connection effect was hypoconnected (or hyperconnected), separately for each disorder (6 contrasts). By accounting for how often a given pair of networks was tested across studies, our procedure thus controlled for the increased number of discoveries merely explained by the larger size of some networks and/or the bias towards a larger interest in some networks in the literature. To test for differences between hypoconnectivity and hyperconnectivity proportions within each disorder (3 contrasts), we employed a similar permutation testing approach, except with a test statistic equal to the absolute difference between the proportions of studies reporting hypoconnectivity and hyperconnectivity effects. Differences in the proportion of hypoconnectivity (or hyperconnectivity) between pairs of disorders (6 contrasts) were tested in the same way, except with a test statistic equal to the absolute difference between the proportion test statistics of 2 disorders.

Permutation testing for 15 contrasts for each of the 78 connections resulted in 1,170 tests. To control the false discovery rate (FDR) of the tests, we employed an empirical Bayes approach that directly modelled the distributions of null and alternative p-values (60). This approach accounted for the atypical distributions of discretely supported p-values generated via Monte-Carlo methods and for the observed positive and negative correlations among p-values, which violate the assumptions of the classical Benjamini-Hocheberg FDR procedure (61). All results reported at the connection level were significant at q^FDR^ < 0.1, this threshold being chosen to best balance the risks of false positives (type I error) and false negatives (type II error), which are respectively problematic for drawing conclusions about disorder-specific and transdiagnostic dysconnectivity patterns.

### Data and code availability

All data as well as Python and R scripts necessary to reproduce the findings reported here are available on Github: https://github.com/pnplab/LBMA. The volumetric version of the CAB-NP atlas can be obtained on Figshare: https://figshare.com/articles/dataset/CAB-NP_projected_on_MNI2009a_GM_volumetric_in_NIfTI_format/14200109.

## RESULTS

### Selected studies

There was some disparity in the extent to which the 3 disorders were investigated in the literature. Of the 428 studies included in our meta-analysis, 37% of them characterized MDD (7,429 patients / 7,414 controls), 19% examined BD (3,330 patients / 4,096 controls) and 52% investigated SZ (11,168 patients / 11,754 controls) (Figure 2A). Most studies (61%) employed a seed-based approach to characterize functional connectivity of regions of interest with the whole brain, at the voxel level (Figure 2B). Critically, a large part (40%) of the selected studies did not report stereotactic coordinates (Figure 2B).

### General network dysconnectivity patterns

At the level of individual studies, 32% of all within- and between-network connections were tested for statistical effects across studies. 25% of those tested connections were reported as significantly impaired, being either hypoconnected or hyperconnected. Similar proportions were observed in MDD, BD and SZ. The distribution of tested and impaired (hypoconnected or hyperconnected) connections across studies was shown to be reproducible when contrasting two independent labels extractions, with Dice similarity coefficients of 0.93 and 0.81 for tested and impaired connections, respectively.

Not all networks, with 12 connections each, were similarly tested (Supplemental Figure S2). The bias towards testing some network connections more than others took a similar form across disorders (r = 0.92-0.97, all ps < 0.001) (Figure 3A, Supplemental Figure S3A). Overall, the more a network connection was tested, the more there was evidence for its impairment while controlling for increased testing of some connections. This was observed for both hypoconnectivity (r = 0.73, p < 0.001) and hyperconnectivity (r = 0.71, p < 0.001) patterns, similarly in all 3 disorders (r = 0.61-0.72, all ps < 0.001) (Figure 3B, Supplemental Figure S3B). The general trend was that the amount of evidence for hypoconnectivity or hyperconnectivity varied across network connections similarly for MDD, BD and SZ (r = 0.77-0.86, all ps < 0.001) (Figure 3B, Supplemental Figure S3B). Across disorders and connections, there was more evidence for hypoconnectivity than hyperconnectivity overall (t = 8.10, p < 0.001). Yet, somewhat paradoxically, connections with larger evidence for hypoconnectivity were also those with more demonstration of hyperconnectivity (r = 0.88, p < 0.001), again similarly in all 3 disorders (r = 0.69-0.88, all ps < 0.001) (Figure 3B, Supplemental Figure S3B).

**Figure 3.**
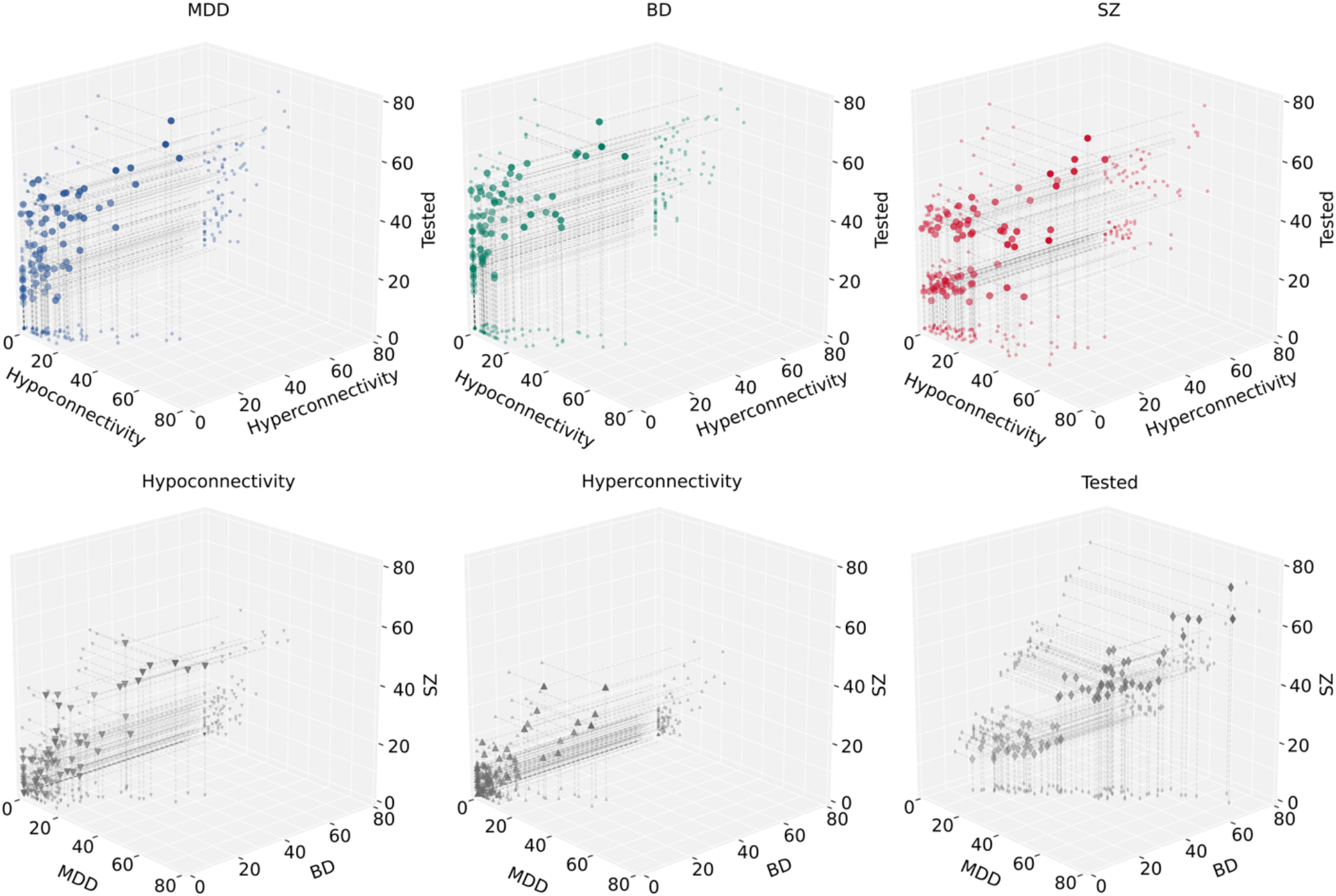
General network dysconnectivity patterns across psychiatric disorders are depicted in 3D correlations plots. In each psychiatric disorder, there were marked positive correlations between how often the 78 connections between pairs of networks were tested for a statistical effect across studies, how often there was a report of hypoconnectivity among those tested connections, and how often hyperconnectivity was shown (upper row). The proportions (%) of how often the 78 connections were tested or reported as functionally impaired (hypoconnectivity or hyperconnectivity) were strongly positively correlated among psychiatric disorders (lower row). MDD, major depressive disorder; BP, bipolar disorder; SZ, schizophrenia.

### Network dysconnectivity among higher-order networks

All within- and between-network connections among the FPN, CON and DMN showed both significant hypoconnectivity and hyperconnectivity in all 3 disorders (all significant results reported hereafter survived a q^FDR^ < 0.1 threshold). While there was a consistent trend towards more evidence of hypoconnectivity than hyperconnectivity for all those connections across disorders, this effect was only significant for 4 out of 6 connections (excluding DMN-DMN and FPN-DMN) in SZ (Figure 4). The consistent trend towards more evidence of hypoconnectivity among the FPN, CON and DMN networks in SZ compared to MDD and BD was not significant for any within- or between-network connection. Direct pairwise comparisons between disorders did not reveal any significant difference in the amount of evidence for either hypoconnectivity or hyperconnectivity among these higher-order networks (Figure 5). It should however be noted that exploratory analysis at the region rather than network level, as reported here, suggests that between-disorders differences may emerge once considering dysconnectivity between small brain regions rather than large-scale brain networks (Supplemental Figure S4).

**Figure 4.**
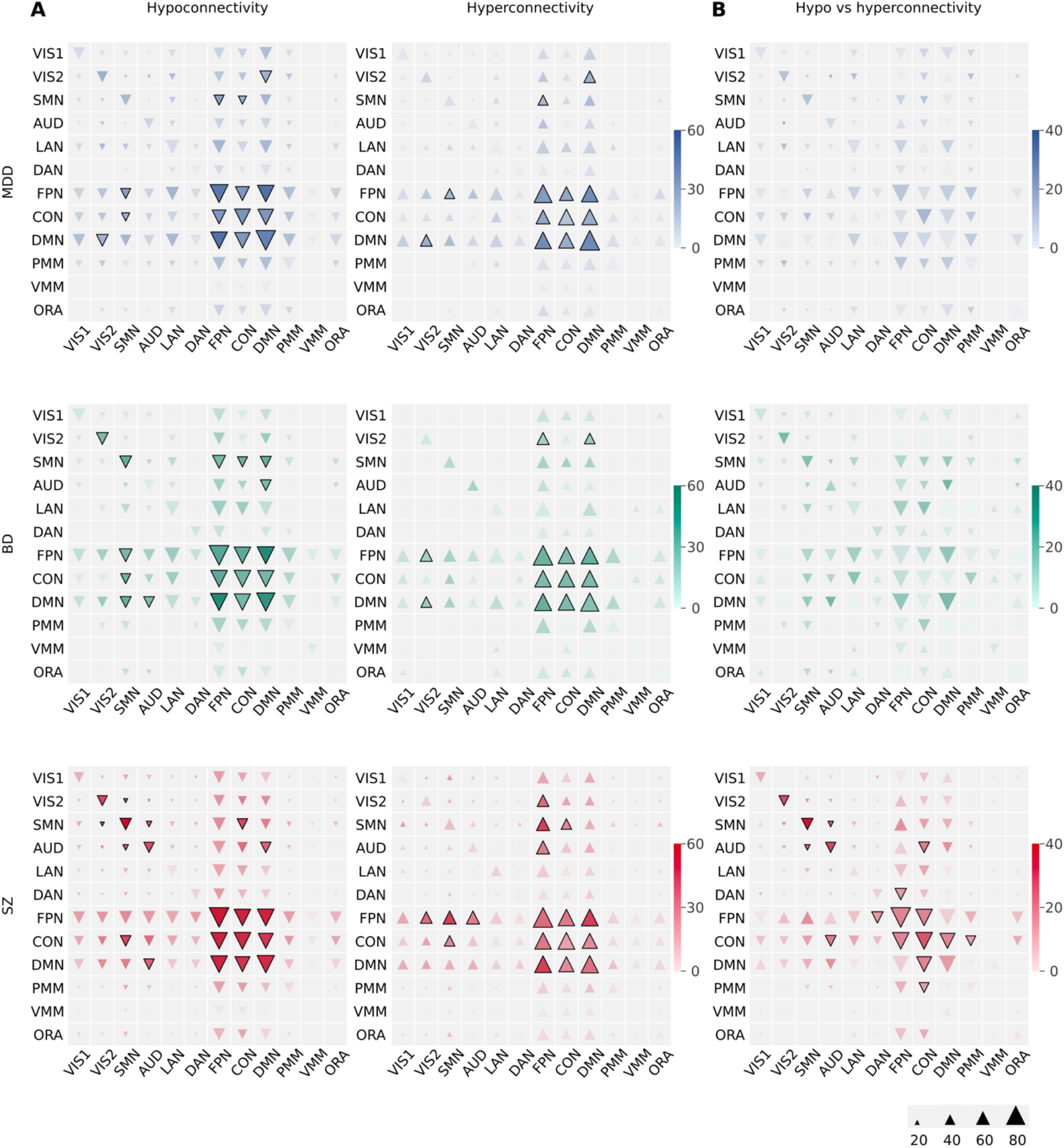
Network dysconnectivity in each psychiatric disorder (MDD, major depressive disorder, blue; BP, bipolar disorder, green; SZ, schizophrenia, red) is shown separately for hypoconnectivity (down-pointing triangles) and hyperconnectivity (up-pointing triangles) effects. Indication of more evidence for hypoconnectivity than hyperconnectivity, and conversely, is shown separately. The size of triangles reflects the proportion (%) of studies that conducted a statistical test for each of the 78 connections. The colors darkness indicates the proportion of studies (%) that reported a statistical effect, relative to how often a connection was tested. Thick edges around triangles represent dysconnectivity effects that were significant at q^FDR^ < 0.1.

**Figure 5.**
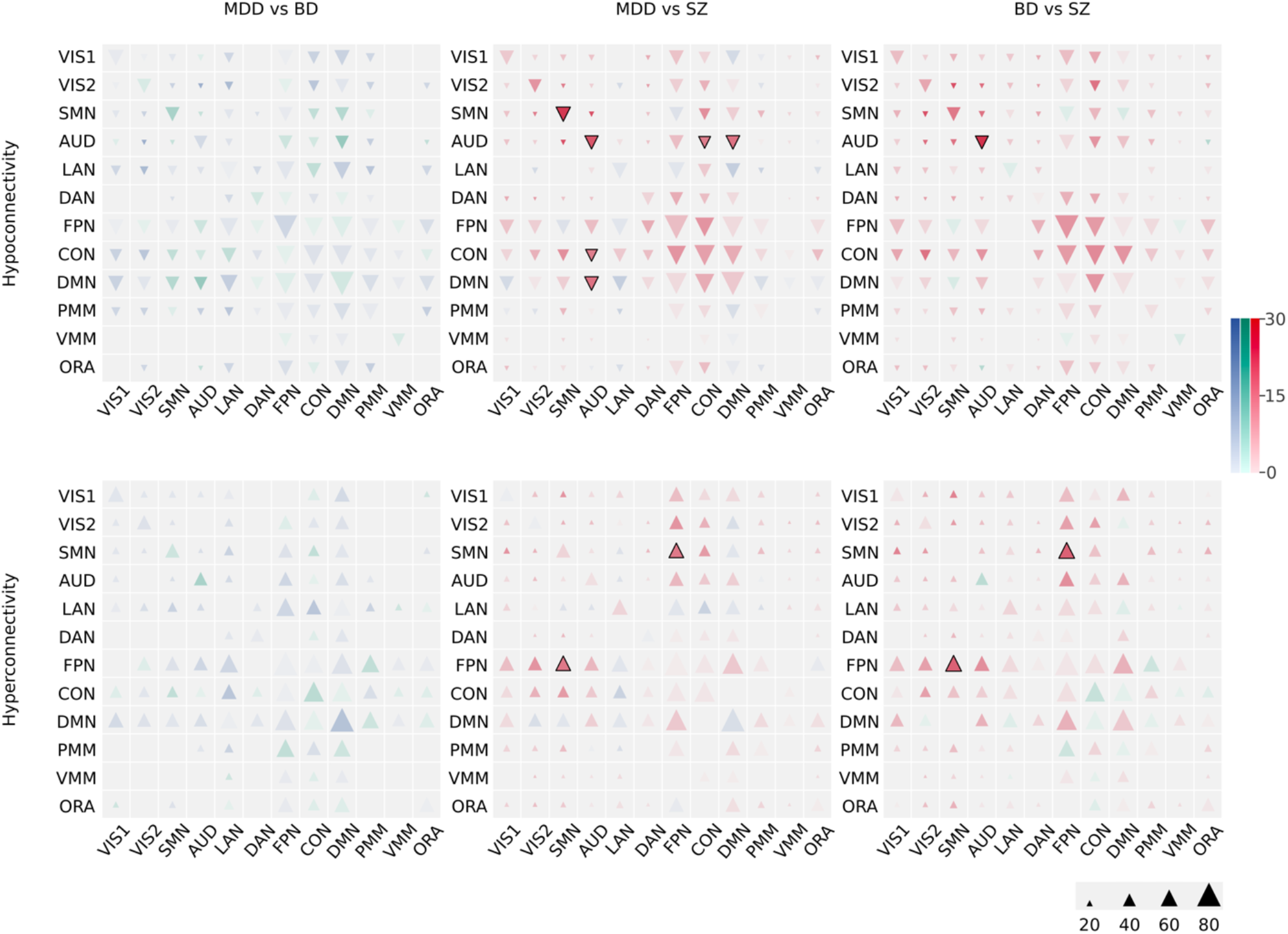
Network dysconnectivity differences between psychiatric disorders (MDD, major depressive disorder, blue; BP, bipolar disorder, green; SZ, schizophrenia, red) is shown separately for greater hypoconnectivity (down-pointing triangles) and greater hyperconnectivity (up-pointing triangles) effects in one disorder than another. The size of triangles reflects the proportion (%) of studies (averaged between disorders) that conducted a statistical test for each of the 78 connections. The colors darkness indicates the between-disorders difference in the proportion of studies (%) that reported a statistical effect, relative to how often a connection was tested (on average in 2 disorders). Thick edges around triangles represent differences in dysconnectivity effects that were significant at q^FDR^ < 0.1.

### Network dysconnectivity of lower-order networks

Lower-order networks showed impaired connectivity with some of the above higher-order networks, either transdiagnostically or with significant differences between disorders (Figure 4). Regarding the SMN, SMN-CON hypoconnectivity was observed in all 3 disorders, yet with SMN-CON hyperconnectivity being also evidenced in SZ. SMN-FPN hypoconnectivity was seen in MDD and BD, while SMN-FPN hyperconnectivity was observed in MDD and SZ. Noteworthy, SMN-FPN hyperconnectivity was more observed in SZ than MDD and BD. SMN-DMN hypoconnectivity was only seen in BD. Regarding VIS2, VIS2-FPN hyperconnectivity was evidenced in both BD and SZ while VIS2-DMN hyperconnectivity was seen in MDD and SZ, yet with VIS2-DMN hypoconnectivity being also observed in MDD. Finally, regarding AUD, AUD-DMN hypoconnectivity was observed in both BD and SZ, with more evidence of AUD-DMN in SZ than MDD. AUD-CON, for which hypoconnectivity was more observed than hyperconnectivity in SZ only, was more hypoconnected in SZ than MDD. AUD-FPN hyperconnectivity was only seen in SZ.

Among lower-order networks, there was evidence of shared SMN-SMN and VIS2-VIS2 hypoconnectivity in BD and SZ, but not MDD (Figure 4). SMN-VIS2, SMN-AUD and AUD-AUD hypoconnectivity was only observed in SZ. There was evidence of more hypoconnectivity than hyperconnectivity for SMN-SMN, VIS2-VIS2, AUD-AUD and SMN-AUD in SZ. Pairwise comparisons between disorders indicated that there is more evidence for AUD-AUD hypoconnectivity in SZ compared to both MDD and BD and SMN-SMN hypoconnectivity in SZ compared to MDD (Figure 5).

## DISCUSSION

Meta-analytic findings reveal similar functional brain dysconnectivity within and between the FPN, CON and DMN networks across mood and psychotic disorders, suggesting higher-order network dysconnectivity is mostly transdiagnostic in nature at the large-scale network level. By contrast, dysconnectivity patterns within lower-order networks such as the SMN and AUD networks as well as between these lower-order networks and higher-order networks were shown to differ between disorders, notably differentiating SZ from BD and MDD. Consistent dysconnectivity patterns were not evidenced for other networks such as LAN and ORA networks.

### Higher-order network dysconnectivity

Highly significant evidence for dysconnectivity patterns shared by all three disorders (MDD, BD, SZ) among heteromodal networks implicating the prefrontal cortex (FPN, CON, DMN) reflect their key role in current models of psychopathology (5,7,8). The various cognitive functions supported by these networks, such as executive control and self-referential monitoring (62–65), are indeed impaired across mood and psychotic disorders (66–68). Accordingly, both original (31,33,35,69) and meta-analytical (13,42,43) transdiagnostic works highlight shared functional brain connectivity abnormalities of the FPN, CON and DMN across a wide range of psychiatric disorders or in relation to a general psychopathology factor (16,25).

Both increased and decreased abnormal connectivity was evidenced for each neurocognitive network, although hypoconnectivity was more frequently reported, particularly in SZ. This paradoxical result might not only be explained by inconsistencies in the literature but also by our choice to explore dysconnectivity at the large-scale network rather than region level. Distinct regions that compose a network are likely to be characterized by opposite dysconnectivity patterns. For instance, the DMN has been reported to be hypoconnected to the ventral insula but hyperconnected to the dorsal insula across several psychiatric disorders (13). This spatial granularity issue may similarly account for the lack of significant differences in the dysconnectivity of neurocognitive networks between MDD, BD and SZ in this study. Our targeted analysis conducted at the region level and some past studies that reported findings at the region or voxel level (12,14,31,32,52) lend support to this hypothesis.

### Lower-order network dysconnectivity

Strong evidence that the functional brain connectivity of unimodal networks (SMN, VIS, AUD) is impaired in both mood and psychotic disorders might seem surprising, given it is seldomly the focus of psychiatric brain imaging. The present meta-analytical results nonetheless indicate that connectivity alterations within motor and sensory networks as well as between them and neurocognitive networks are often reported, hence underscoring a lack of emphasis on such findings in the literature. This observation echoes recent calls to better promote research centered on motor and sensory systems in psychiatric neuroscience, as exemplified by the delayed inclusion of a domain dedicated to motor systems (70,71) and the suggestion to add a sensory processing domain (72) in the Research Domain Criteria (RDoC) framework (73). Motor abnormalities that include neurological soft signs, extrapyramidal symptoms (dyskinesia, parkinsonism) and catatonic phenomena, are observed in a wide range of disorders (74,75). Similarly, aberrant sensory processing and perceptual signaling are encountered in disorders other than SZ (72,76,77). Accordingly, we observed several dysconnectivity patterns of motor (SMN) and sensory (VIS2, AUD) networks being shared by at least two disorders, in line with previous studies that reported transdiagnostic alterations of unimodal networks using fMRI (26,27,32,33,78). A notable result was however that, in some instances, there was more evidence for dysconnectivity of these networks (SMN, AUD) in SZ compared to BD and MDD. Gradients of impairments in connectivity that scale as a function of illness severity along the mood/psychosis continuum have been reported before (35,37,78), and may account for more aggravated motor symptoms (74) and the frequent presence of auditory hallucinations in SZ (79).

### Strengths and limitations

The main strength of the present study lies in the use of text labels rather than stereotactic coordinates as the source of information for our meta-analysis (44–46). By doing so, a comprehensive inclusion of numerous prior studies that did not report coordinates was possible. Moreover, dysconnectivity patterns could be explored for the whole brain rather than focussing on a few selected networks of interest. The use of a whole-brain cortico-subcortical atlas that includes atypical auditory and language networks (59) further represents an improvement over previous meta-analyses. Besides, the manual extraction of text labels and subsequent automatic assignment to large-scale networks were shown to be reproducible. A future step will be to apply natural language processing algorithms to fully automatize the extraction of relevant papers and text labels (80,81), making our label-based meta-analytical approach an even more appealing alternative to traditional (coordinate-based) meta-analyses of brain imaging findings.

The systematic exploration of whole-brain connectivity comes at a cost. Multiple testing over all connections with sufficient statistical power can only be performed for a limited number of brain networks, not dozens or hundreds of local brain regions given the amount of available published studies. As aforementioned, networks such as the DMN or CON merge together distinct brain areas or subnetworks with distinct connectivity profiles and functions (64,82), and are thus likely to be differentially impacted by psychopathology (12,13). In addition, we adopted a conservative approach where a brain area corresponding to a text label could only be assigned to a single network, which it maximally overlapped with. The complexity of small regions with multimodal integration zones that in fact belong to multiple networks (e.g., the thalamus (83)) was thus ignored. For the same reason, the inferior frontal gyrus and amygdala were not respectively assigned to the LAN and ORA in our analysis, as one would have expected (84,85). This may account for the lack of significant findings in these networks, despite past evidence of their roles in language disturbances associated with thought disorder and verbal hallucinations in SZ (85,86) or emotional dysregulation across mood and psychotic disorders (84,87,88). As additional research in the field accumulates, we are hopeful that future label-based meta-analyses will be able to provide finer-grained assessments of functional brain dysconnectivity in psychiatric disorders.

## Conclusions

Using a novel meta-analytical approach, we explored the relatedness of functional brain dysconnectivity patterns across 3 major psychiatric disorders. In line with prevailing models of psychopathology, transdiagnostic abnormal connectivity among core higher-order neurocognitive networks was highlighted. More surprisingly, lower-order (motor, visual, auditory) networks were also affected across disorders, however revealing gradients of impairment from mood to psychotic disorders. These findings underscore the role that motor and sensory processes play in the etiology of major psychiatric disorders and thus call for dedicated research on this topic (71,72).

## Data Availability

All data as well as Python and R scripts necessary to reproduce the findings reported here are available on Github: https://github.com/pnplab/LBMA. The volumetric version of the CAB-NP atlas can be obtained on Figshare: https://figshare.com/articles/dataset/CAB- NP_projected_on_MNI2009a_GM_volumetric_in_NIfTI_format/14200109.

## ACKNOWLDEGMENTS AND DISCLOSURES

This work was funded by a salary award “chercheur boursier junior 1” of the “Fonds de recherche du Québec – Santé” [to PO], a Canadian Institutes of Health Research (CIHR) project grant (Grant No. PJT-165910 [to PO]), and the Courtois foundation through the Courtois NeuroMod Project (https://www.cneuromod.ca) [to PO]. SP is holder of the Eli Lilly Canada Chair on schizophrenia research.

SG1 and PO were responsible for study concept and design. SG1, SS, SP, MD, VP and PO were responsible for acquisition, analysis, or interpretation of data. SS, HN and PO were responsible for statistical analysis. SG1, HN and PO were responsible for drafting of the manuscript. SP, SG2 were responsible for critical revision of the manuscript for important intellectual content. PO obtained funding and was responsible for study supervision.

The authors report no biomedical financial interests or potential conflicts of interest.

## ARTICLE INFORMATION

From the Research Center of the Montreal University Institute for Mental Health (SG1, SS, SP, MD, VP, PO) and Department of Psychiatry and Addictology, University of Montreal (SG1, SP, PO), Montréal, Québec, Canada; and the Institute of Neuroscience and Medicine, Brain and Behavior (INM-7), Research Centre Jülich, Jülich and Institute of Systems Neuroscience, Medical Faculty, Heinrich-Heine-University Düsseldorf, Düsseldorf, Germany (SG2); and the School of Mathematics and Physics, University of Queensland, St. Lucia, Queensland and Department of Mathematics and Statistics, Latrobe University, Melbourne, Victoria, Australia (HN).

SG1 and SS contributed equally to this work as joint first authors.

## SUPPLEMENTAL INFORMATION

### SUPPLEMENTAL METHODS

Description of permutation testing method For each disorder *k* ∈ 𝒟 = {MDD,BD,SZ}, we define the tested connections via the array 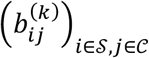, where 𝒮 = {1, …, *S*} and 𝒞 = {1, …, *C*} are the indices of the investigated studies and connections, respectively. Here, 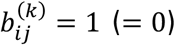 indicates that connection *j* was investigated (was not investigated) in study *j*, with respect to disorder *k*. For each *k*, we further define the set of indices for which tested connections were observed: 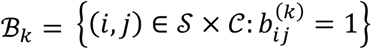, and define the array 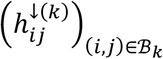 where 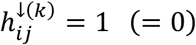 if hypoconnectivity was detected (was not detected) in study *i*, for connection *j*. Similarly, we define the array 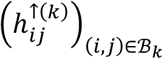, where 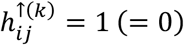 if hyperconnectivity was detected (was not detected) in study *i*, for connection *j*.

Using these data, we wish to test three types of hypotheses, for each pair of connection and disorder (*j, k*)∈ 𝒞 × 𝒟. ***Hypotheses of type A1 and A2:*** For pair (*j, k*), is there evidence of hypoconnectivity. For pair (*j, k*), is there evidence of hyperconnectivity. ***Hypotheses of type B:*** For pair (*j, k*), is there evidence that the proportions of studies reporting hyper and hypoconnectivity differ. Lastly, we consider for distinct triples (*j, k, l*)∈ 𝒞 × 𝒟 × 𝒟, the following hypotheses: ***Hypotheses of type C1 and C2:*** For triple (***j, k, l***), is there evidence that the proportions of studies reporting hypoconnectivity differ between disorder ***k*** and disorder ***l***. For triple (***j, k, l***), is there evidence that the proportions of studies reporting hyperconnectivity differ between disorder ***k*** and disorder ***l***.

In order to test the hypotheses above, we employ permutation-based tests (Monte Carlo tests), via the independence decomposition framework of Zhu (2005) (1). By Definition 1.2.1 of Zhu (2005) (1), we say that a random vector **x** is independently decomposable if **x** = **Az**, in distribution, where **A** (an operator; e.g. a matrix) and **z** (a vector) are independent.

Suppose that we observe data (**x**_1_, …, **x**_*N*_. Using these data, we can test the hypotheses H_0_: **x** is independently decomposable versus H_1_: **x** is not independently decomposable. To do so, we construct some test statistic *T*_0_=*τ*(**x**_1_, …, **x**_*N*_), which equals in distribution to *T* =*τ*(**A**_1_**z**_1_, …, **A**_*N*_**z**_*N*_), under the null hypothesis. To assess whether the null hypothesis is true, we simulate *M* independent instances of *T*, by computing 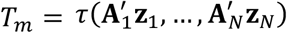, where 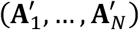has the same distribution as (**A**_1_, …, **A**_*N*_). This simulation process can be thought of as a Monte-Carlo simulation. When **A**_1_, …, **A**_*N*_ are functions of randomly sampled permutation matrices, then the test is usually referred to as a permutation test.

The *p*-value associated with test statistic *T*_0_ can be computed as

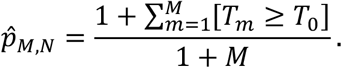

By Proposition 1.2.1 of Zhu (2005) (1), we have the fact that for any *α*∈ (0, 1):

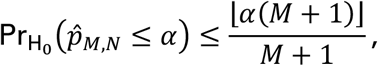

where ⌊*a*⌋ is the integer part of *a*, and where 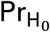 is the probability measure under H_0_. See Lehmann and Romano (2005; Sec. 15.2) (2) for additional details.

#### Hypotheses of type A1 and A2

For tests of type A1, for disorder *k* and connection *j*, we identify 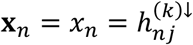, where *n* ∈ {*i*: (*i, j*) ∈ ℬ_*k*_} and *N* = #{*i*: (*i, j*) ∈ ℬ_*k*_}. Under the null hypothesis, we assume the independent decomposition **x**_*n*_ = [**A**_*n*_]_1•_**z**_*n*_, where **A**_*n*_ is a uniform randomly sampled permutation of the set {*j* ′: (*n, j*′) ∈ ℬ_*k*_}, [**A**_*n*_]_1•_ is the first row of **A**_*n*_, and 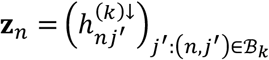. That is, under the null hypothesis, for each study *n* under which a tested connection was observed for connection 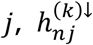 is assumed to be equal to 1 with probability proportional to the number of hypoconnections observed for study *n*, as a proportion to the number of tested connections observed in study *n*.

We wish to test that there is an increased level of hypoconnectivity for connection *j*. To do so, we define the observed test statistic

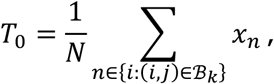

and simulate *M* statistics *T*_*m*_ under the null hypothesis, with form

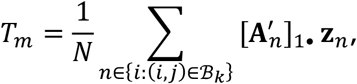

where 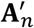 is uniform randomly generated with the same distribution as **A**_*n*_.

The test statistic *T*_0_ is just the proportion of studies that identified that connection *j* was hypoconnected, out of the studies for which a tested connection was observed for *j*. The simulated values *T*_*m*_ are then simulations of this proportion, under the probability model described above. Thus, the obtained *p*-value 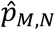, can be read as the proportion of test statistics {*T*_0_, …, *T*_*m*_} that were greater or equal to the observed statistic *T*_0_. That is, the proportion of simulations where the number of studies that identified connection *j* as being hypoconnected was greater than the observed number of hypoconnected studies.

For tests of type A2, we may modify the description above by replacing the symbol ↓ by ↑, and the prefix hypo by hyper.

#### Hypotheses of type B

For tests of type B, for disorder *k* and connection *j*, we start by defining the vector concatenation

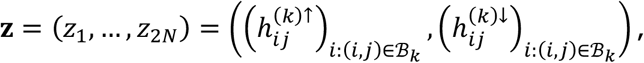

where **z** is a vector of length 2*N* with *N* = #{*i*: (*i, j*) ∈ ℬ_*k*_}. We also observe as data **x** = **z**. Under the null hypothesis, we assume the independent decomposition **x** = **Az**, where **A** is uniform random permutation matrix corresponding to a permutation of the positions of **z**. That is, under the null hypothesis, the probability of any study (with observed tested connection) being observed as hypo or hyperconnected at connection *j* is equal, with pooled probability equal to

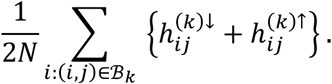

We wish to test that the hypo and hyperconnectivity proportions for connection *j* are different. To do so, we define the observed test statistic

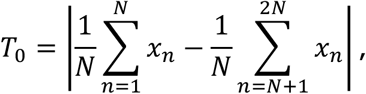

and simulate *M* statistics *T*_*m*_ under the null hypothesis, with form

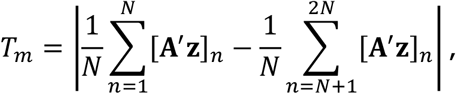

where **A**′ is uniform randomly generated with the same distribution as **A**, and [**A**′ **z**]_*n*_ is the *n*th element of the vector **A**′ **z**.

The test statistic *T*_0_ is just the absolute difference between the proportions of studies that identified that connection *j* was hypoconnected versus hyperconnected, out of the studies for which a tested connection was observed for *j*. The simulated values *T*_*m*_ are then simulations of this difference in proportion, under the probability model described above. The *p*-value 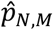 can then be interpreted as the proportion of simulations where the difference between hypo and hyperconnected studies for connection *j* was greater than the observed difference in proportion.

#### Hypotheses of type C1 and C2

For tests of type C1, for disorder *k* and *l*, and connection *j*, we start by defining the vector concatenation

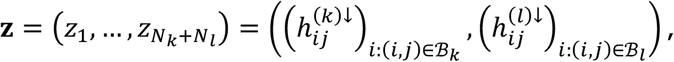

where **z** is a vector of length *N*_*k*_ + *N*_*l*_ with *N*_*k*_ = #{*i*: (*i, j*) ∈ ℬ_*k*_} and *N*_*l*_ = #{*i*: (*i, j*) ∈ ℬ_*l*_}. We also observe as data **x** = **z**. Under the null hypothesis, we assume the independent decomposition **x** = **Az**, where **A** is uniform random permutation matrix corresponding to a permutation of the positions of **z**. That is, under the null hypothesis, the probability of any study across both disorders (with observed tested connection) being observed as hypoconnected at connection *j* is equal, with pooled probability equal to

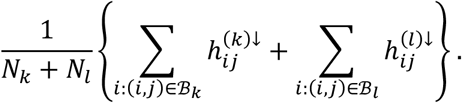

We wish to test that the proportions of hypoconnected studies for connection *j* are different between disorders *k* and *l*. To do so, we define the observed test statistic

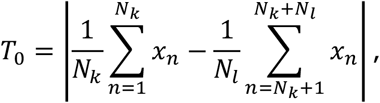

and simulate *M* statistics *T*_*m*_ under the null hypothesis, with form

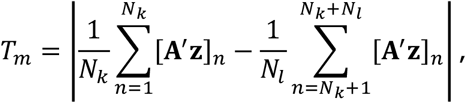

where **A**′ is uniform randomly generated with the same distribution as **A**.

The test statistic *T*_0_ is the absolute difference between the proportions of studies that identified that connection *j* was hypoconnected for disorders *k* and *l*. The simulated values *T*_*m*_ are then simulations of this difference in proportion, under the probability model described above. The *p*-value 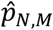 can then be interpreted as the proportion of simulations where the difference between proportions of studies that identified hypoconnectivity for connection *j* for disorders *k* and *l* was greater than the observed difference in proportion.

For tests of type C2, we may modify the description above by replacing the symbol ↓ by ↑, and the prefix hypo by hyper.

#### False discovery rate control

Assume that we observe *N p*-values *p*_1_, …, *p*_*N*_ in (0, 1). We then convert the *p*-values via the probit transformation to obtain *z*-values:

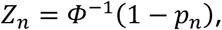

where ***Φ***^− 1^ is the inverse cumulative distribution function of the standard normal distribution, and *n* ∈ {1, …, *N*}. The empirical Bayes of McLachlan et al. (2006) (3) and Efron (2010) (4) assumes that, marginally, *z*_*n*_ has mixture distribution defined by the density function

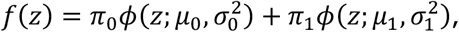

where *π*_0_, *π*_1_ ∈ (0, 1) with *π*_0_ + *π*_1_ = 1, and where *ϕ*(.; *μ, σ*^2^) is the normal density function with mean *μ* and variance *σ*^2^. Here, *π*_0_ can be interpreted as the marginal probability that a *z*-value corresponds a test that for which the null hypothesis is true. Densities 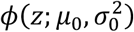 and 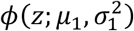 are thus the densities of the *z*-values, conditional on the null hypothesis of the underlying test being true or false, respectively. Since we would assume that smaller *p*-values provide evidence of false null hypotheses, it is assumed that *μ*_1_ > *μ*_0_.

Notice that under the typical theory of well-specified tests, *p*_*n*_ ∼ Uniform(0, 1) if *p*_*n*_ the null hypothesis is true, and thus *p*_*n*_ ∼ Normal(0, 1). Efron (2010) (4) provides compelling argument that Uniform(0, 1) is often not the distribution of true null *p*-values in practice, when tests may be misspecified or are non-standard, and argues for the use of the so-called empirical null density 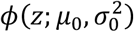, where *μ*_0_≈ 0 and 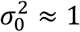. We will also make use of this flexibility, since it is clear that our own *p*-values are non-standard, due to the use of permutation testing.

For known parameters 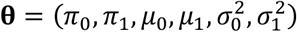, suppose that we reject hypothesis *n* if *τ*_*n*_(**θ**) ≤ *κ* and *z*_*n*_ > *μ*_0_. for some *κ* ∈ [0, 1], where

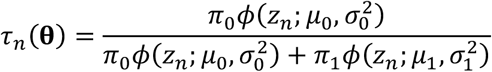

is the a posteriori probability that hypothesis *n* has a true null, conditioned on observation of its *z*-value *z*_*n*_. If we use the aforementioned rejection rule across all *N* hypotheses, then we can estimate the false discovery rate (FDR) due to this rule via the expression

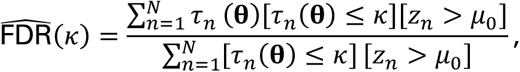

which is a consistent estimate of the FDR under so-called *M*-dependence (cf. Nguyen et al. 2014 (5), Thm. 1). We define the *q*-value (FDR adjusted *p*-value) *q*_*n*_ for each hypothesis *n* as the estimated FDR given that *κ* =*τ*_*n*_(**θ**):

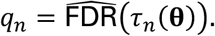

In practical applications, we must estimate **θ** by some 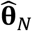, which is obtained via the procedure described in Nguyen et al. (2019) (6).

### Secondary analysis at the region level within high-order networks

A secondary analysis was conducted at the region level, focussing only on regions that maximally overlapped with either the fronto-parietal (FPN), cingulo-opercular (CON) or default-mode (DMN) network from the Cole-Anticevic Brain Network Partition (CAB-NP) atlas (1). Translation of the labels manually extracted at various spatial resolutions into the AAL3 space (7) and coding the presence or absence of tested, hypoconnected or hyperconnected connections were accomplished using the same procedure as described for the main analyses. Hemispheric dissociation was ignored, thus leading to investigate 48 brain regions. Of the 48 within-region and 1128 between-region connections, only the top 3 connections with the larger between-disorder dysconnectivity effects (either hypo or hyperconnectivity) were reported for each of the 6 within and between-network connections among higher-order networks (Supplemental Figure S4).

## SUPPLEMENTAL TABLES

**Table S1:** Brief definitions of the methodological approaches included in the meta-analysis.

| Method | Definition | Nature of connectivity |
| --- | --- | --- |
| Seed-based voxel-wise (SBVW) | Correlation between a seed and every voxel of the brain. | Distant |
| Seed-to-region (STR) | Correlation between a seed and a specific brain region or network. | Distant |
| Network-based connectome-wide (NBCW) | Pairwise correlations between every brain network. | Distant |
| Independent component analysis (ICA) | Temporally correlated voxels grouped into multiple independent networks. | Distant |
| Voxel-mirrored homotopic connectivity (VMHC) | Correlation between pairs of symmetric inter-hemispheric voxels. | Inter-hemispheric |
| Regional homogeneity (ReHo) | Correlation of a voxel's timeseries with its neighboring voxels. | Local |
| Amplitude of low-frequency fluctuation (ALFF) | Total power of the BOLD signal within the low frequencies range (0.01 and 0.1 Hz). | Local |

**Table S2:** Network sizes defined by their proportions (%) of the total volume of the Cole-Anticevic Brain Network Partition (CAB-NP) atlas (1).

| **Network name** | **size** |
| --- | --- |
| Primary visual network (VIS1) | 4.54 % |
| Secondary visual network (VIS2) | 11.99 % |
| Somatomotor network (SMN) | 13.03 % |
| Auditory network (AUD) | 2.06 % |
| Language network (LAN) | 5.25 % |
| Dorsal attentional network (DAN) | 5.82 % |
| Frontoparietal network (FPN) | 17.53 % |
| Cingulo-opercular network (CON) | 14.13 % |
| Default-mode network (DMN) | 21.01 % |
| Posterior-multimodal network (PMN) | 1.76 % |
| Ventral-multimodal network (VMN) | 1.87 % |
| Orbito-affective network (ORA) | 1.02 % |

VIS1, primary visual network; VIS2, secondary visual network; SMN, sensorimotor network; AUD, auditory network; LAN, language network; DAN, dorsal attentional network; FPN, fronto-parietal network; CON, cingulo-opercular network; DMN, default-mode network; PMN, posterior multimodal network; VMN, ventral multimodal network; ORA, orbito-affective network.

## SUPPLEMENTAL FIGURES

**Figure S1:**
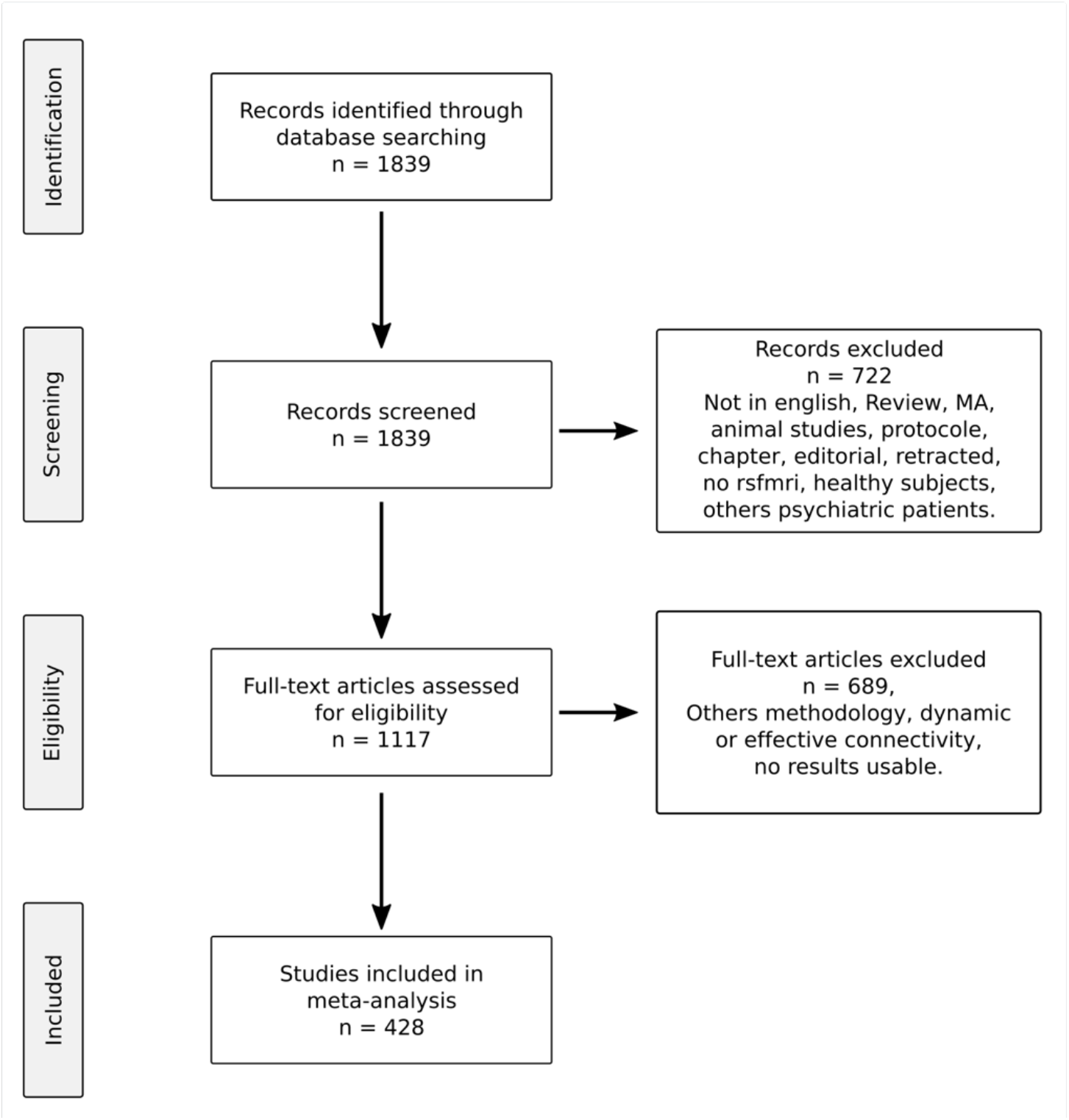
Selection process of the 428 studies included in the meta-analysis is presented in a flowchart according to the Preferred Reporting Items for Systematic Reviews and Meta-Analyses (PRISMA) guidelines. MA: meta-analysis; rsfmri: resting-state functional magnetic resonance imaging.

**Figure S2:**
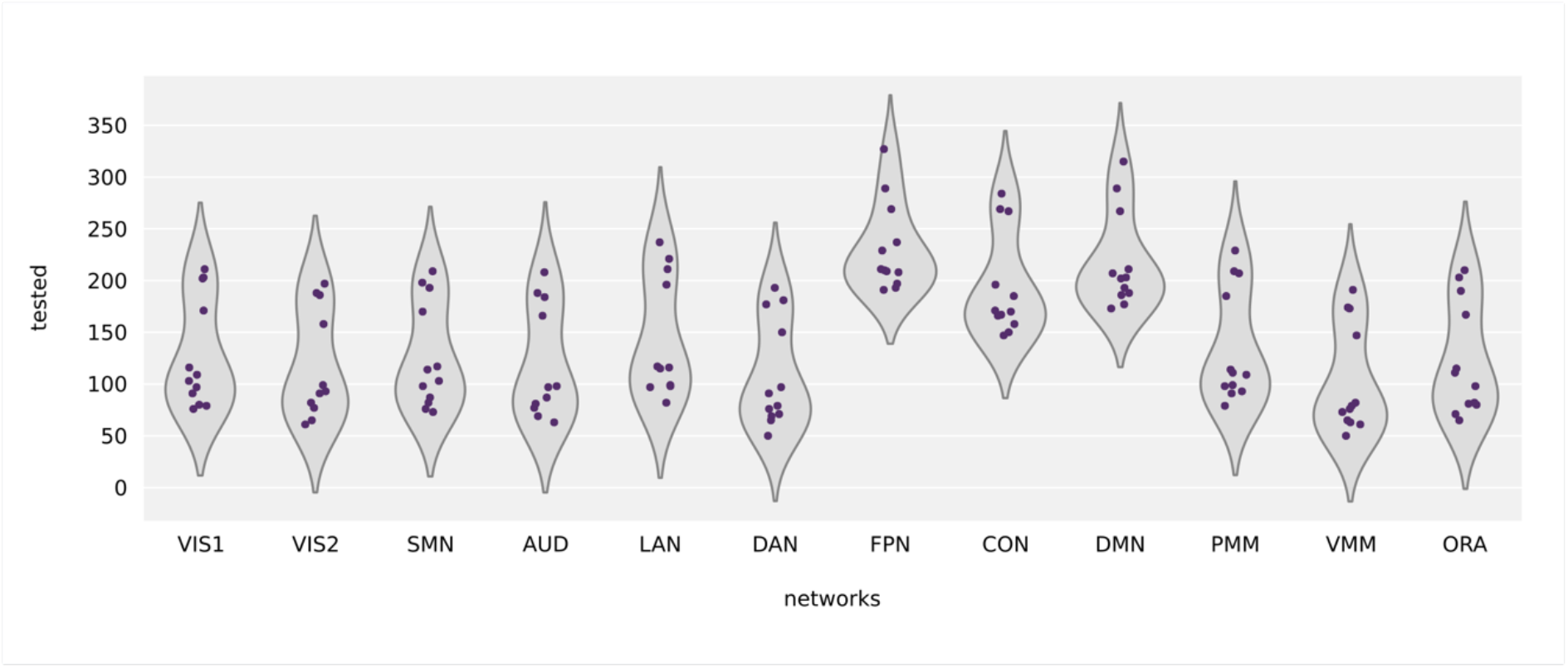
Distribution of the number of studies that conducted a statistical test by connections across psychiatric disorders is shown separately for each network. The 12 connections between a specific network and all the CAB-NP atlas networks are depicted in each violin plot. VIS1, primary visual network; VIS2, secondary visual network; SMN, sensorimotor network; AUD, auditory network; LAN, language network; DAN, dorsal attentional network; FPN, fronto-parietal network; CON, cingulo-opercular network; DMN, default-mode network; PMN, posterior multimodal network; VMN, ventral multimodal network; ORA, orbito-affective network.

**Figure S3:**
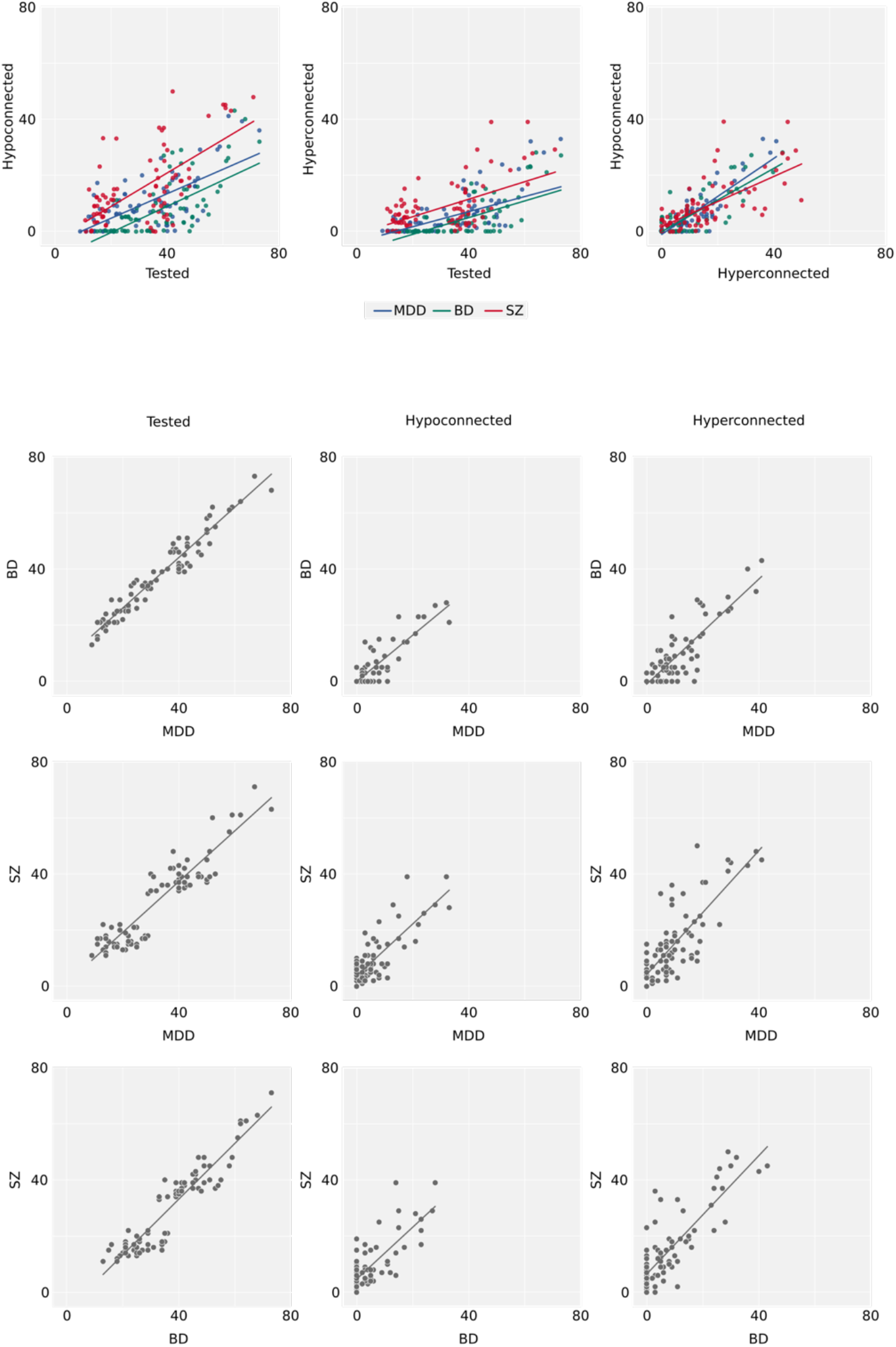
Network dysconnectivity patterns in each psychiatric disorder are depicted in correlations plots. The proportions (%) of how often the 78 connections were tested, how often there was a report of hypoconnectivity and hyperconnectivity among those tested connections were similarly positively correlated in each disorder (MDD, major depressive disorder; BP, bipolar disorder; SZ, schizophrenia). The proportions (%) of how often the 78 connections were tested, hypoconnected or hyperconnected between disorders are shown by pairwise correlations.

**Figure S4:**
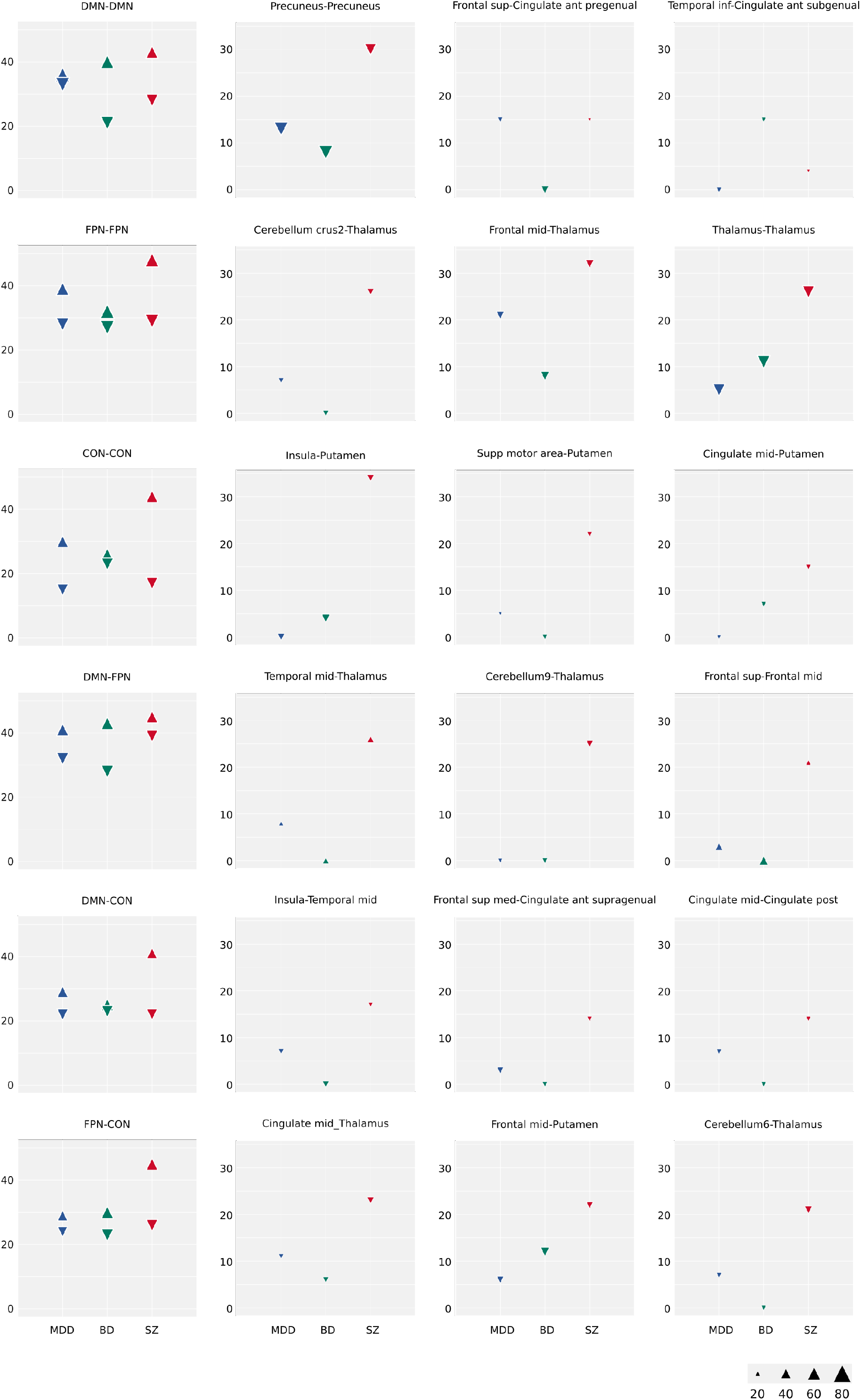
Higher-order networks (FPN, fronto-parietal network; CON, cingulo-opercular network; DMN, default-mode network) hypoconnectivity (down-pointing triangles) and hyperconnectivity (up-pointing triangles) for each psychiatric disorders (MDD, major depressive disorder, blue; BP, bipolar disorder, green; SZ, schizophrenia, red) are shown in the first column. The 3 regional connections with the with the largest between disorder dysconnectivity (either hypoconnectivity or hyperconnectivity) are shown in the row of the corresponding network connection. The size of triangles reflects the proportion (%) of studies that conducted a statistical test for the specified connections. Sup, superior; mid, middle; inf, inferior; ant, anterior; med, median; post, posterior; supp, supplementary.

